# Environmental heavy metal exposure, socioeconomic inequalities, and childhood anaemia in Bangladesh: a nationally representative, survey-weighted Bayesian hierarchical analysis

**DOI:** 10.64898/2026.09.13.26362929

**Authors:** Md Fuad Al Fidah, Md Ridwan Islam, Anjan Kumar Roy, Rubhana Raqib, Tahmeed Ahmed, Mustafa Mahfuz

**Affiliations:** Nutrition Research Division, icddr,b, Mohakhali, Dhaka; Office of the Executive Director, icddr,b, Mohakhali, Dhaka

**Keywords:** Anemia, Child, Heavy Metals, Arsenic, Cadmium, Mercury, Socioeconomic Factors, Bangladesh, Bayesian Analysis, Environmental Health

## Abstract

**Background:** Childhood anaemia remains a major public health challenge in low- and middle-income countries. Although nutritional deficiencies are well-established contributors, the population-level role of environmental heavy metal exposure remains poorly understood in South Asia.

**Objectives:** To estimate the prevalence, socioeconomic inequalities, and factors associated with childhood anaemia among children aged 12-59 months in Bangladesh, with particular emphasis on environmental heavy metal exposure.

**Methods:** We analysed nationally representative cross-sectional data from the Bangladesh Multiple Indicator Cluster Survey (MICS) 2025, including 10,343 children aged 12-59 months. Blood concentrations of lead, arsenic, cadmium, and mercury were transformed using base-2 logarithms (log_2_). The primary outcome was binary child anaemia status. Survey-weighted Bayesian hierarchical logistic regression using R-INLA, with primary sampling unit (PSU)-level random intercepts, was the primary analytical approach. Socioeconomic inequalities were evaluated using the Slope Index of Inequality (SII) and Relative Index of Inequality (RII). Population attributable fractions (PAFs) were estimated separately using counterfactual analyses.

**Results:** Overall, 4779 (44.3%) children had anaemia. In multiple Bayesian models, each 2-fold increase in blood arsenic (posterior aOR 1.12, 95% CrI 1.06-1.18; P(aOR>1): >0.99) and blood cadmium (aOR 1.09, 95% CrI 1.04-1.13; P(aOR>1): >0.99) was independently associated with higher odds of childhood anaemia. Conversely, higher blood mercury was inversely associated (aOR 0.82, 95% CrI 0.78-0.86; P(aOR<1): >0.99). Older child age (aOR 0.69 per 13.2 months increase, 95% CrI 0.65-0.72), higher Height-for-Age Z-score (aOR 0.88, 95% CrI 0.84-0.91), maternal higher education (aOR 0.69, 95% CrI 0.56-0.86), and richest wealth status (aOR 0.80, 95% CrI 0.66-0.97) were associated with lower anaemia odds. Anaemia was more common among children from the poorest households (SII: −12.39%, 95% CI −17.62, −7.17), while anaemia prevalence was lower among wealthier households (RII: 0.76, 95% CI 0.67-0.85).

**Interpretation:** Childhood anaemia in Bangladesh is associated with environmental toxicant exposures, nutritional deficits, and structural socioeconomic disadvantage. The findings highlight the potential importance of integrating nutritional, environmental, and socioeconomic considerations within strategies to address the high burden of child anaemia in Bangladesh.

## Introduction

Anaemia remains one of the most pervasive global public health challenges, disproportionately burdening young children in low- and middle-income countries (LMICs).^1^ Globally, an estimated 39.8% of children under five years of age are affected by anaemia, with the highest concentration observed in South Asia and sub-Saharan Africa.^2,3^ In early childhood, particularly among children aged 12–59 months who are undergoing rapid somatic growth and neurodevelopment, adequate haemoglobin concentrations are vital for cellular oxygen transport. Chronic or severe anaemia during this critical developmental window is associated with irreversible cognitive impairment, compromised motor development, blunted immune function, and elevated mortality.^4^ Consequently, reducing the prevalence of childhood anaemia is a core priority of the World Health Organization’s Global Nutrition Targets and Sustainable Development Goal 2.2.^4^

Historically, public health strategies to combat childhood anaemia have focused primarily on nutritional deficiencies, infections, and poor infant and young child feeding practices.^2,3^ However, growing evidence suggests that environmental toxicant exposure may also contribute to haematological abnormalities. Studies from Bangladesh have documented substantial childhood exposure to arsenic and lead,^5^ while population-based evidence has linked arsenic exposure with lower haemoglobin concentrations.^6,7^ Heavy metals may affect erythropoiesis through distinct biological pathways. Lead inhibits key enzymes involved in haem synthesis and may impair erythrocyte survival.^8^ Cadmium may disrupt iron homeostasis through interactions with intestinal metal transport pathways, including DMT1, and may contribute to anaemia through renal and oxidative mechanisms.^9^ Arsenic may impair erythropoiesis through oxidative stress, inflammation, and disruption of erythroid differentiation.^6,9,10^ Mercury may interfere with the haematopoietic system by impairing iron metabolism and inducing oxidative stress, which may shorten erythrocyte lifespan ^11^.

Despite the toxicological plausibility linking heavy metal exposure to impaired erythropoiesis, major evidence gaps persist in LMICs. Previous studies investigating environmental heavy metals and childhood health in Bangladesh have been constrained by small sample sizes, localised geographic footprints (e.g., single-district cohorts), or an exclusive focus on a single heavy metal rather than simultaneous multi-element toxicity.^5,12^ Furthermore, studies evaluating how environmental heavy metal burdens interact with persistent socioeconomic inequalities—such as household wealth, maternal education, and water, sanitation, and hygiene (WASH) infrastructure—are scarce to shape population-level anaemia risk.^13^ The release of the Bangladesh Multiple Indicator Cluster Survey (MICS) 2025 provides a landmark opportunity to address these gaps, representing the first nationally representative survey in Bangladesh to incorporate simultaneous blood testing for lead, arsenic, cadmium, and mercury alongside haemoglobin measurement in children aged 12-59 months.^13^ Therefore, this study aimed to evaluate the multi-sectoral factors associated with anaemia among children aged 12-59 months in Bangladesh using the nationally representative MICS 2025 dataset. Specifically, we examined associations of blood heavy metal concentrations, household socioeconomic inequalities, WASH characteristics, and individual demographic determinants with child anaemia.

## Methods

### Data Source and Study Design

This study is a secondary analysis of nationally representative, cross-sectional survey data from the Bangladesh Multiple Indicator Cluster Survey (MICS) 2025. The survey was implemented between February 24 and June 30, 2025, by the Bangladesh Bureau of Statistics (BBS) under the Statistics and Informatics Division (SID), Ministry of Planning, with technical and financial support from UNICEF Bangladesh.^13^ icddr,b Laboratory performed the Laboratory analysis of biomarkers in whole blood. Data collection was conducted using Computer-Assisted Personal Interviewing (CAPI) on CSPro version 7.7.3 installed on Android tablets equipped with Bluetooth communication protocols for real-time field data synchronization.^13^

### Sampling Framework and Participant Selection

The sampling frame for MICS 2025 was derived from the 2022 Bangladesh Population and Housing Census. A multi-stage, stratified cluster sampling design was used to produce statistically reliable estimates at national, urban/rural, divisional (8 administrative divisions), and domain levels (64 districts plus Dhaka North City Corporation, Dhaka South City Corporation, and Chattogram City Corporation, forming 67 main sampling domains).^13^

In the first stage, census Enumeration Areas (EAs)—defined as Primary Sampling Units (PSUs)—were selected within each urban and rural stratum using systematic probability proportional to size (PPS) sampling. In the second stage, following a complete household mapping and listing operation conducted in late 2024, a systematic random sample of 20 households was drawn from each selected EA. In total, 3,149 sample clusters (862 urban and 2,287 rural) and 62,980 households were selected nationwide.

For the blood biomarker testing component, households with children aged 12-59 months were identified. Where more than one eligible child lived in a sampled household, a Kish grid selection module embedded in the CAPI application was used to randomly select one child for blood sample collection. Out of 24,680 under-five children enumerated in the survey, 17,132 children aged 12-59 months were selected for phlebotomy; 10,936 mothers or caregivers provided informed consent (63.8% consent rate). After excluding participants with missing data for haemoglobin, anthropometry, heavy metal measurements or other variables (n = 593), the final analytical sample for our study comprised 10,343 children aged 12-59 months (figure 1).^13^

**Figure 1.**
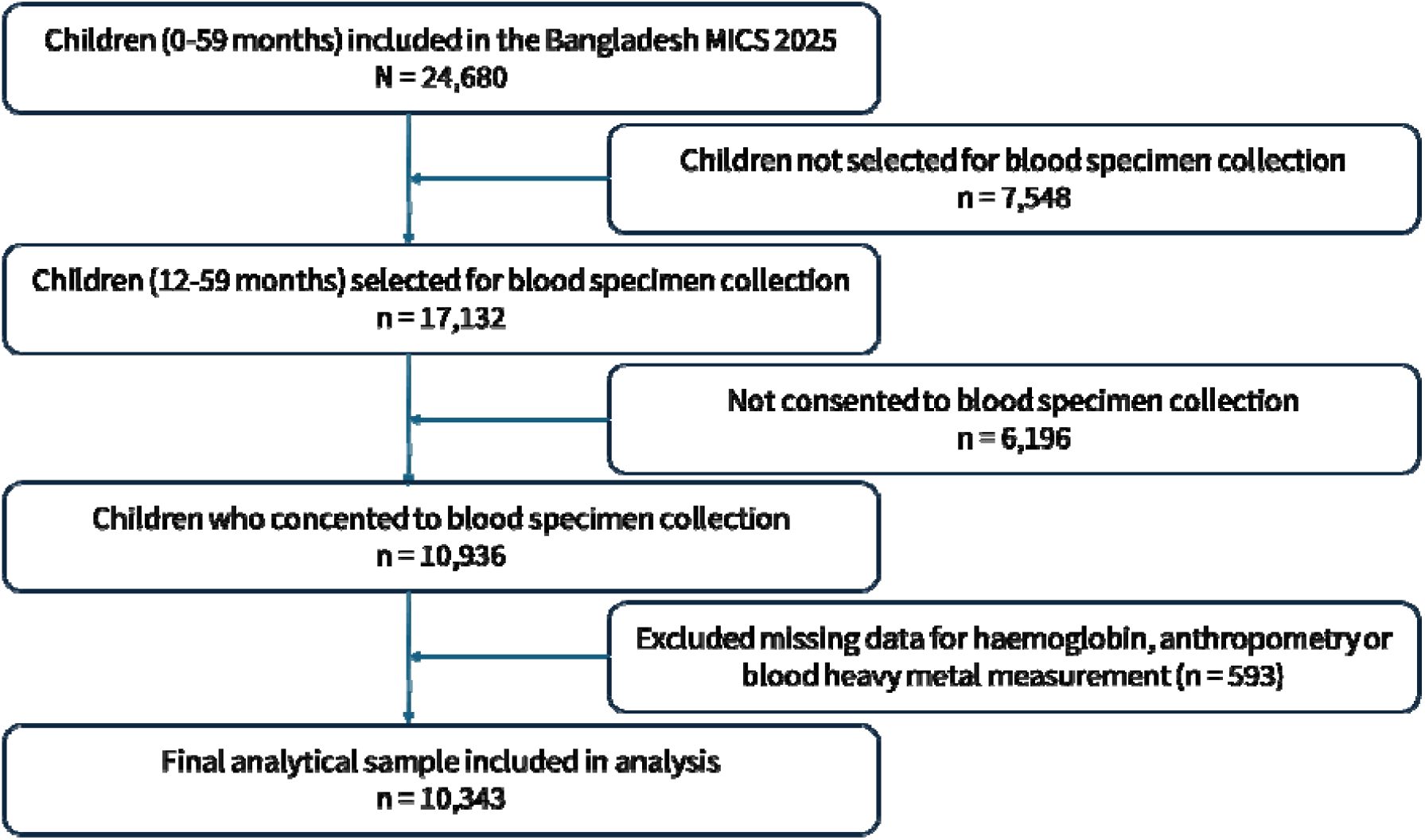
Study participant selection.

### Blood Specimen Collection and Biomarker Measurements

Blood collection was conducted in household settings by 43 dedicated, professionally trained phlebotomists using aseptic precautions and sterile, disposable syringes and needles.^13^ Venous blood specimens (2 ml) were collected into metal free tube by trained phlebotomists and transported to designated reference laboratory in Dhaka specifically the Immunobiology, Nutrition and Toxicology laboratory in icddr,b, where haemoglobin and heavy metal concentrations were measured using validated methods. Concentrations of heavy metals were measured in whole blood using Inductively Coupled Plasma Mass Spectrometry (ICP-MS; NexION® 2000, PerkinElmer) and haemoglobin using the cyanmethemoglobin method. For ICP-MS method, three levels of commercial control ClinChek were used to ensure the accuracy and precision. For haemoglobin assessment, internal pooled whole blood was used as internal quality control.^14^

### Haemoglobin

Haemoglobin was measured in whole blood using cyanmethemoglobin method. Whole blood was mixed with Drabkin’s solution for thirty minutes followed by reading of optical density (OD) using a UV-Vis Spectrophotometers (Evolution 220, Thermo Fisher Scientific, USA) at a wavelength of 540 nm. Three-point calibration curve was prepared using Cyanmethemoglobin Standard Solution and result was calculated based on that curve. Commercial quality control blood (Eightcheck-3WP, Sysmex, Germany) was used to monitor and verify the performance of haemoglobin measurement.

### Heavy metal analysis by ICP-MS

Heavy metals (Lead, Cadmium, Mercury, Arsenic) were simultaneously measured by inductively coupled plasma mass spectrometer (ICP-MS) (NexION 2200, PerkinElmer, USA). Blood samples, including standard and control were digested with concentrated nitric acid. Following digestion, the samples were prepared by adding diluent (mixture of methanol, nitric acid, gold standard and type-1 water) and mixture of internal standards. A set of digested blood based working standards, controls (RECIPE Chemicals + Instruments, GmbH) and diluted digested samples were introduced into the ICP-MS under optimal spectroscopic conditions. Each heavy metal concentration is calculated based on the respective standard curve which was prepared using that standards. The blanks were used to verify the absence of elements carryover and confirm low background levels and three levels of ClinChek Controls (RECIPE Chemicals + Instruments, GmbH) were used to ensure both accuracy and precision of daily laboratory results.

Blood concentrations were quantified for four heavy metals: lead (measured in µg/dL); arsenic (measured in µg/L), cadmium (measured in µg/L), and mercury (measured in µg/L). While the official Survey Findings Report deferred estimates for arsenic, cadmium, and mercury in whole blood due to analytical concerns, these biomarkers reflect different exposure windows. They may not adequately represent cumulative body burden. Whole-blood arsenic may not adequately reflect overall arsenic exposure. Blood cadmium and mercury primarily reflect recent or ongoing exposure rather than cumulative body burden. This study utilised the raw biomonitoring data to investigate underlying public health associations; findings for these exposures should therefore be interpreted in light of these analytical and biomarker limitations. We employed base-2 logarithmic transformations to address right-skewness and used Bayesian hierarchical modelling with Primary Sampling Unit (PSU) random intercepts to specifically account for the spatial heterogeneity and clustering inherent in these environmental exposures. Consequently, a 1-unit increase in transformed concentration corresponds directly to a 2-fold increase (doubling) in raw exposure level across all four biomarkers in multiple regression models.

### Operational Definitions of Variables

#### Operational definition of childhood anaemia

The primary outcome variable was childhood anaemia, defined as a binary variable based on haemoglobin concentration measured in grams per decilitre (g/dL). In accordance with the age-specific haemoglobin cut-offs used in the Bangladesh MICS 2025 and the current WHO definition, children aged 12-23 months with a haemoglobin concentration of <10.5 g/dL and children aged 24-59 months with a haemoglobin concentration of <11.0 g/dL were classified as anaemic (coded as 1).Children with haemoglobin concentrations at or above the respective age-specific cut-off were classified as non-anaemic (coded as 0).^15^

Key independent exposures were blood concentrations of lead, arsenic, cadmium, and mercury. Sociodemographic, household, and health-related covariates included child characteristics like age in months (continuous), sex (male/female), Height-for-Age z-score (HAZ, continuous), acute respiratory infection (ARI) in the preceding 2 weeks (present/absent), and diarrhoea in the preceding 2 weeks (present/absent); maternal characteristics: age in years (continuous) and educational level (up-to primary/upto secondary/higher secondary or more); and household and environmental characteristics like wealth index (poorest/poor/middle/rich/richest), place of residence (urban/rural), administrative division (8 divisions), household overcrowding (>3 persons per sleeping room: present/absent), sanitation facility (improved/unimproved), and handwashing facility (no service, limited service, basic service).^1,2,5,16^

In this study, ARI was treated as a binary variable. Children were coded as 1 if they had ARI and 0 if they did not. ARI was defined based on the mother’s report of a child experiencing a cough accompanied by rapid breathing, short and quick breaths, or difficulty breathing during the two weeks preceding the survey. Children whose symptoms were limited to a blocked nose were not classified as having ARI.^13,17^

The household wealth index was used as an indicator of socioeconomic status. It is a composite measure derived from household ownership of selected assets, housing characteristics, and access to basic services and amenities. The index is constructed using principal component analysis (PCA), which assigns weights to household assets and generates standardized wealth scores with a mean of 0 and a standard deviation of 1.^18^ Based on these scores, households are ranked and classified into five wealth quintiles: poorest, poorer, middle, richer, and richest.^13,17,19^

Sanitation facility was classified according to the WHO/UNICEF Joint Monitoring Programme (JMP) classification. Households using flush or pour-flush toilets connected to a piped sewer system, septic tank, pit latrine, or covered drain; ventilated improved pit latrines; pit latrines with slabs; twin pit latrines with slabs; composting toilets; or container-based sanitation systems were classified as improved (coded as 1). Households using flush or pour-flush toilets discharging into an open drain or an unknown destination, pit latrines without a slab, bucket toilets, hanging toilets/latrines, other unimproved sanitation facilities, or having no sanitation facility (open defecation) were classified as having an unimproved (coded as 0).^20^

Handwashing facility was classified according to the WHO/UNICEF JMP. Households were categorised as having basic service (coded as 2) if a handwashing facility was observed with both water and soap or detergent available at the time of the survey. Households with an observed handwashing facility but lacking either water or soap/detergent were classified as having a limited service (coded as 1). Households with no handwashing facility observed were classified as having no service (coded as 0).^21^

Overcrowding was defined as a binary variable based on the number of household members per sleeping room. The overcrowding ratio was calculated by dividing the total number of household members by the number of sleeping rooms in the household. Households with more than three persons per sleeping room were classified as present (coded as 1), whereas those with three or fewer persons per sleeping room were classified as absent (coded as 0).^22^

### Statistical Analysis

Data preparation and variable recoding were conducted using Stata version 19.0 (StataCorp LLC, College Station, TX, USA) and jamovi (version 2.7). All inferential statistical modeling, survey-weighted estimations, Bayesian hierarchical regressions, model diagnostics, predictive calibration, and health inequality analyses were executed in R version 4.4.3 (R Foundation for Statistical Computing, Vienna, Austria) utilizing RStudio (Posit Software, PBC).

To represent the national target population of children aged 12-59 months, all analyses accounted for the multi-stage stratified cluster sampling design. Individual blood sampling weights were normalized to sum to the analytical complete-case sample size (N = 10,343), ensuring accurate likelihood computation in Bayesian and frequentist models. Blood heavy metal concentrations (lead, arsenic, cadmium, and mercury) exhibited pronounced right-skewness and were transformed using base-2 logarithms (log2). Consequently, a 1-unit increase in transformed concentration corresponds directly to a doubling (2-fold increase) in raw exposure level.

Continuous demographic covariates like child age (SD = 13.2 months) and maternal age (SD = 6.3 years), were standardized to Z-scores (Mean = 0, SD = 1) to enable standardised comparison of effect sizes. Height-for-Age Z-score (HAZ) was retained on its native WHO growth standard scale (1 unit = +1.0 WHO Z-score). Categorical covariates were formatted as factors with pre-specified reference categories. To account for the multi-stage stratified cluster sampling design and non-response, survey weights were incorporated into all analyses specifying primary sampling units (psu), strata, and individual child’s blood sampling weights for Survey-weighted descriptive and frequentist regression analyses.^13^

Primary multivariable associations between heavy metal exposures and child anaemia were evaluated using survey-weighted Bayesian hierarchical logistic regression performed via Integrated Nested Laplace Approximations (INLA) in the R-INLA package.^23,24^ Primary sampling unit (PSU)-level clustering was accommodated by incorporating random intercepts, f(psu, model = “iid”), while administrative division and place of residence were included as covariates. To preserve computational neutrality without imposing rigid parametric constraints, weakly informative Normal priors centered at zero were specified: β ~ Normal(0, SD = 1.5) [Precision = 0.444] for all fixed-effect regression slopes, and α ~ Normal(0, SD = 2.5) [Precision = 0.160] for the global intercept.^25,26^ Inference was based on posterior median adjusted Odds Ratios (posterior aORs) accompanied by 95% Bayesian Credible Intervals (95% CrIs).

To evaluate directional certainty independent of p-value thresholds, directional posterior probabilities P(aOR > 1) and P(aOR < 1) were computed using marginal posterior distribution functions. Evidence strength was categorized as very strong (P≥ 0.99), strong (0.95≤P<0.99), moderate (0.90≤P<0.95), or weak (P<0.90).^27^ Model goodness-of-fit was formally evaluated using the Deviance Information Criterion (DIC)^28^ and Widely Applicable Information Criterion (WAIC).^29^ The full multiple model was compared with an intercept-only null model, with ΔDIC > 10 and ΔWAIC > 10 indicating substantially better model fit. Cross-validatory predictive performance was assessed using the Conditional Predictive Ordinate (CPO), with the number of CPO failures reported as an indicator of numerical or predictive instability.

To test analytical robustness against frequentist paradigms, a conventional survey-weighted multiple logistic regression model was fitted using the survey R package (svyglm with quasibinomial variance structure). Goodness-of-fit for the frequentist model was evaluated using the Archer-Lemeshow calibration test for complex survey design.^30^ To test the validity of assuming log-linear dose–response relationships in the primary model, we conducted sensitivity analyses evaluating non-linearity between blood heavy metal concentrations and childhood anaemia. First, each heavy metal was modelled using survey-weighted multivariable logistic regression with restricted cubic splines, specifying four knots positioned at standard percentiles (5th, 35th, 65th, and 95th).^31^ Design-adjusted Wald tests were performed to formally test for departures from linearity (evaluating the joint significance of the second and third spline terms). Second, to aid clinical interpretation and evaluate dose-dependent threshold effects where non-linearity was identified, blood lead concentration was additionally categorized according to international benchmark thresholds: <3.5 μg/dL,^32^ 3.5 to <5.0 μg/dL, 5.0 to <10.0 μg/dL,^33^ and ≥10.0 μg/dL.

Absolute and relative socioeconomic inequalities in child anaemia across household wealth quintiles were quantified using the Slope Index of Inequality (SII) and Relative Index of Inequality (RII), estimated using the ‘healthequal’ package in R according to the World Health Organization Health Equity Monitor framework.^34^

PAFs for selected exposures and risk factors were estimated using posterior counterfactual predictions from multivariable Bayesian logistic regression models fitted using INLA. For each factor, the observed dataset was combined with a counterfactual dataset. All individuals were assigned to the specified target, while other covariates remained at their observed values. A joint model was then fitted to the observed and counterfactual datasets. For each posterior draw, individual predicted probabilities of anaemia were obtained for both scenarios and aggregated using normalised sampling weights to estimate the observed prevalence, P_observed_, and counterfactual prevalence, P_counterfactual_. The PAF was calculated as: PAF(%)= {(P_observed_ − P_counterfactual_) / P_observed_} ×100

The point estimate was derived from posterior mean predicted probabilities, and 95% CrI were obtained from the PAF distribution across 1,000 posterior samples. For cadmium, arsenic, and mercury, the counterfactual target was exposure at or below the sample median; for blood lead, the target was <5.0 µg/dL.^13^ Other factors were assigned to their respective prespecified target categories. These PAFs represent model-based counterfactual estimates under the specified exposure scenarios and should not be interpreted as direct estimates of the effects of real-world interventions.

### Ethical Considerations

The Bangladesh MICS 2025 survey protocol, including the blood specimen collection procedure, was formally reviewed and approved by the icddr,b Ethical Review Committee in November 2024 (Protocol Reference: PR-24113).^13^ Written informed consent was obtained from mothers or legal guardians prior to blood collection.

Participants identified with severe anaemia or dangerously elevated heavy metal concentrations were provided immediate counselling and referred through the Ministry of Health and Family Welfare to appropriate medical facilities for clinical management.^13^ Anonymized microdata were accessed for research purposes in accordance with BBS and UNICEF data sharing policies.

## Results

Among 10,343 children included in the analysis, 4,779 had anaemia, corresponding to a survey-weighted prevalence of 44.3%. Compared with non-anaemic children, those with anaemia were younger (mean age: 35.1 vs. 38.6 months) and had lower mean height-for-age Z scores (−1.4 vs. −1.2). A lower proportion of anaemic children had mothers with up to higher education (16.7% vs. 21.1%) and belonged to the richest wealth quintile (14.2% vs. 18.4%). However, the median blood mercury level among the anaemic children was lower than non-anaemic children (median: 1.4 vs 1.5 µg /L) (Table 1).

**Table 1.**
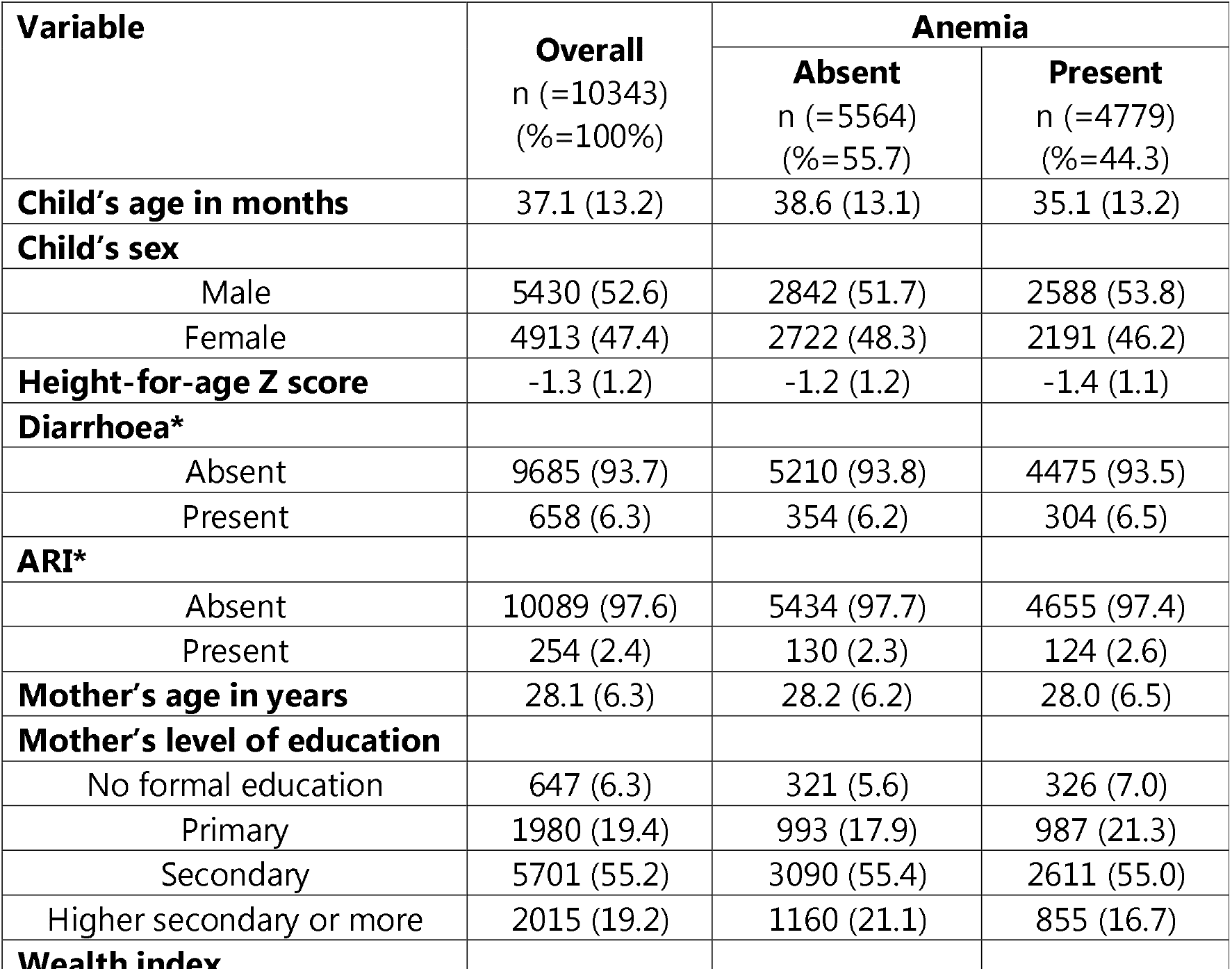
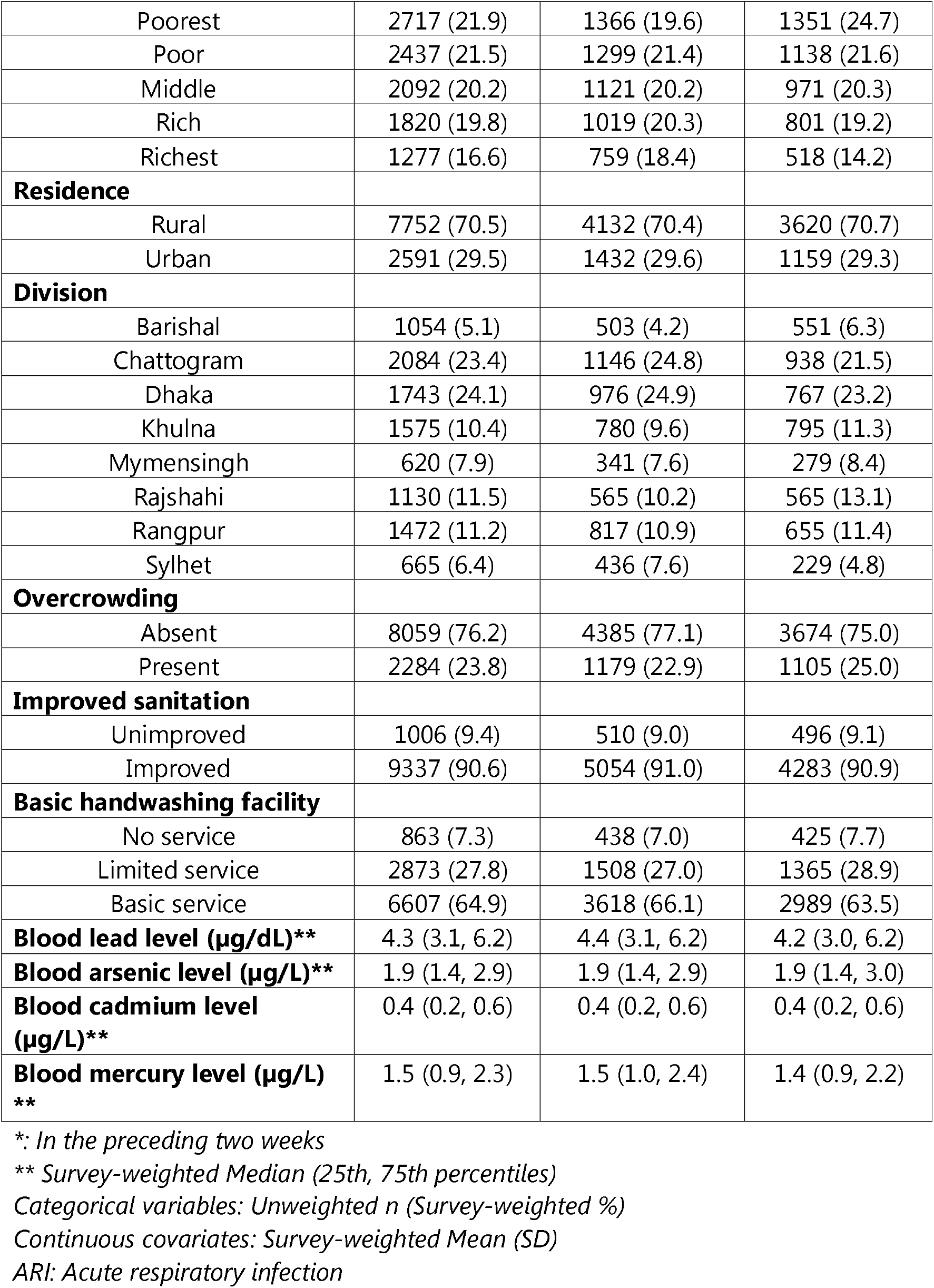
Characteristics of the study participants by childhood anaemia status.

| Variable | Overall<br>n (=10343)<br>(%=100%) | Anemia |  |
| --- | --- | --- | --- |
|  |  | Absent<br>n (=5564)<br>(%=55.7) | Present<br>n (=4779)<br>(%=44.3) |
| **Child's age in months** | 37.1 (13.2) | 38.6 (13.1) | 35.1 (13.2) |
| **Child's sex** |  |  |  |
| Male | 5430 (52.6) | 2842 (51.7) | 2588 (53.8) |
| Female | 4913 (47.4) | 2722 (48.3) | 2191 (46.2) |
| **Height-for-age Z score** | -1.3 (1.2) | -1.2 (1.2) | -1.4 (1.1) |
| **Diarrhoea\*** |  |  |  |
| Absent | 9685 (93.7) | 5210 (93.8) | 4475 (93.5) |
| Present | 658 (6.3) | 354 (6.2) | 304 (6.5) |
| **ARI\*** |  |  |  |
| Absent | 10089 (97.6) | 5434 (97.7) | 4655 (97.4) |
| Present | 254 (2.4) | 130 (2.3) | 124 (2.6) |
| **Mother's age in years** | 28.1 (6.3) | 28.2 (6.2) | 28.0 (6.5) |
| **Mother's level of education** |  |  |  |
| No formal education | 647 (6.3) | 321 (5.6) | 326 (7.0) |
| Primary | 1980 (19.4) | 993 (17.9) | 987 (21.3) |
| Secondary | 5701 (55.2) | 3090 (55.4) | 2611 (55.0) |
| Higher secondary or more | 2015 (19.2) | 1160 (21.1) | 855 (16.7) |
| **Wealth index** |  |  |  |
| Poorest | 2717 (21.9) | 1366 (19.6) | 1351 (24.7) |
| Poor | 2437 (21.5) | 1299 (21.4) | 1138 (21.6) |
| Middle | 2092 (20.2) | 1121 (20.2) | 971 (20.3) |
| Rich | 1820 (19.8) | 1019 (20.3) | 801 (19.2) |
| Richest | 1277 (16.6) | 759 (18.4) | 518 (14.2) |
| **Residence** |  |  |  |
| Rural | 7752 (70.5) | 4132 (70.4) | 3620 (70.7) |
| Urban | 2591 (29.5) | 1432 (29.6) | 1159 (29.3) |
| **Division** |  |  |  |
| Barishal | 1054 (5.1) | 503 (4.2) | 551 (6.3) |
| Chattogram | 2084 (23.4) | 1146 (24.8) | 938 (21.5) |
| Dhaka | 1743 (24.1) | 976 (24.9) | 767 (23.2) |
| Khulna | 1575 (10.4) | 780 (9.6) | 795 (11.3) |
| Mymensingh | 620 (7.9) | 341 (7.6) | 279 (8.4) |
| Rajshahi | 1130 (11.5) | 565 (10.2) | 565 (13.1) |
| Rangpur | 1472 (11.2) | 817 (10.9) | 655 (11.4) |
| Sylhet | 665 (6.4) | 436 (7.6) | 229 (4.8) |
| **Overcrowding** |  |  |  |
| Absent | 8059 (76.2) | 4385 (77.1) | 3674 (75.0) |
| Present | 2284 (23.8) | 1179 (22.9) | 1105 (25.0) |
| **Improved sanitation** |  |  |  |
| Unimproved | 1006 (9.4) | 510 (9.0) | 496 (9.1) |
| Improved | 9337 (90.6) | 5054 (91.0) | 4283 (90.9) |
| **Basic handwashing facility** |  |  |  |
| No service | 863 (7.3) | 438 (7.0) | 425 (7.7) |
| Limited service | 2873 (27.8) | 1508 (27.0) | 1365 (28.9) |
| Basic service | 6607 (64.9) | 3618 (66.1) | 2989 (63.5) |
| **Blood lead level (µg/dL)\*\*** | 4.3 (3.1, 6.2) | 4.4 (3.1, 6.2) | 4.2 (3.0, 6.2) |
| **Blood arsenic level (µg/L)\*\*** | 1.9 (1.4, 2.9) | 1.9 (1.4, 2.9) | 1.9 (1.4, 3.0) |
| **Blood cadmium level (µg/L)\*\*** | 0.4 (0.2, 0.6) | 0.4 (0.2, 0.6) | 0.4 (0.2, 0.6) |
| **Blood mercury level (µg/L)**<br>**\*\*** | 1.5 (0.9, 2.3) | 1.5 (1.0, 2.4) | 1.4 (0.9, 2.2) |

\*: In the preceding two weeks
\*\* Survey-weighted Median (25th, 75th percentiles)
Categorical variables: Unweighted n (Survey-weighted %)
Continuous covariates: Survey-weighted Mean (SD)
ARI: Acute respiratory infection

In the primary survey-weighted Bayesian hierarchical logistic regression model, blood concentrations of arsenic and cadmium were positively associated with elevated odds of child anaemia (Table 2). Each 2-fold increase (doubling) in blood arsenic concentration was associated with 12% higher odds of anaemia (posterior aOR = 1.12, 95% CrI: 1.06-1.18; P(aOR > 1) > 0.99). Similarly, each doubling of blood cadmium was associated with 9% higher odds of anaemia (aOR = 1.09, 95% CrI: 1.04-1.13; P(aOR > 1) > 0.99). Conversely, blood lead (aOR = 0.92, 95% CrI: 0.86-0.98; P(aOR < 1) > 0.99) and mercury (aOR = 0.82, 95% CrI: 0.78-0.86; P(aOR < 1) > 0.99) exhibited inverse associations after adjusting for co-exposures and primary sampling unit (PSU) random intercepts.

**Table 2.**
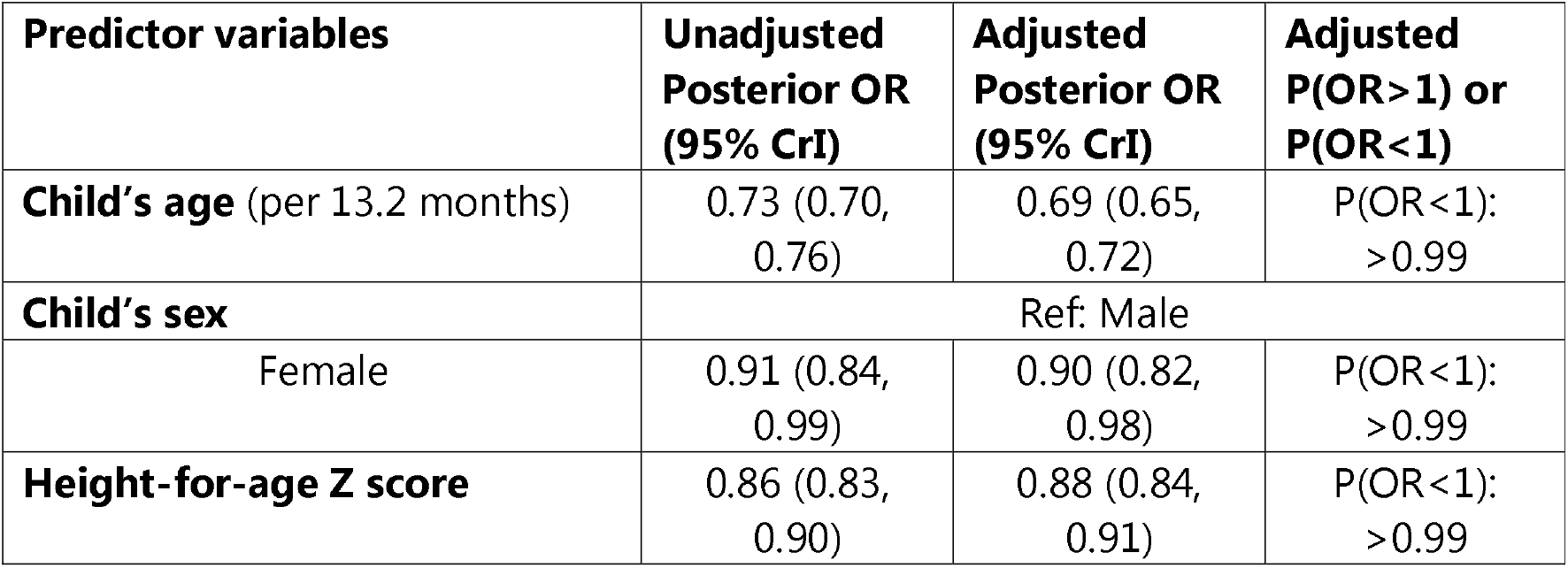
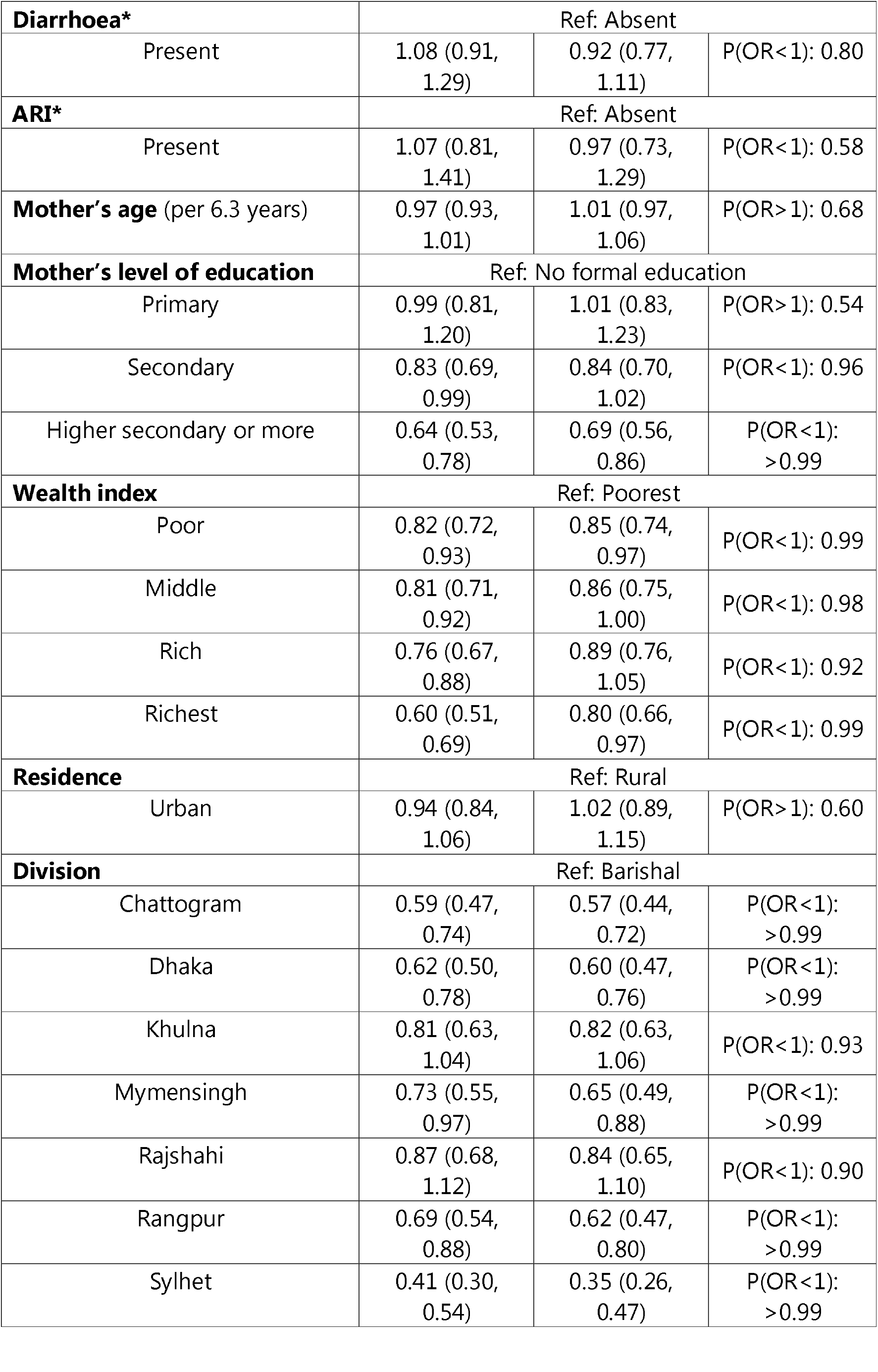
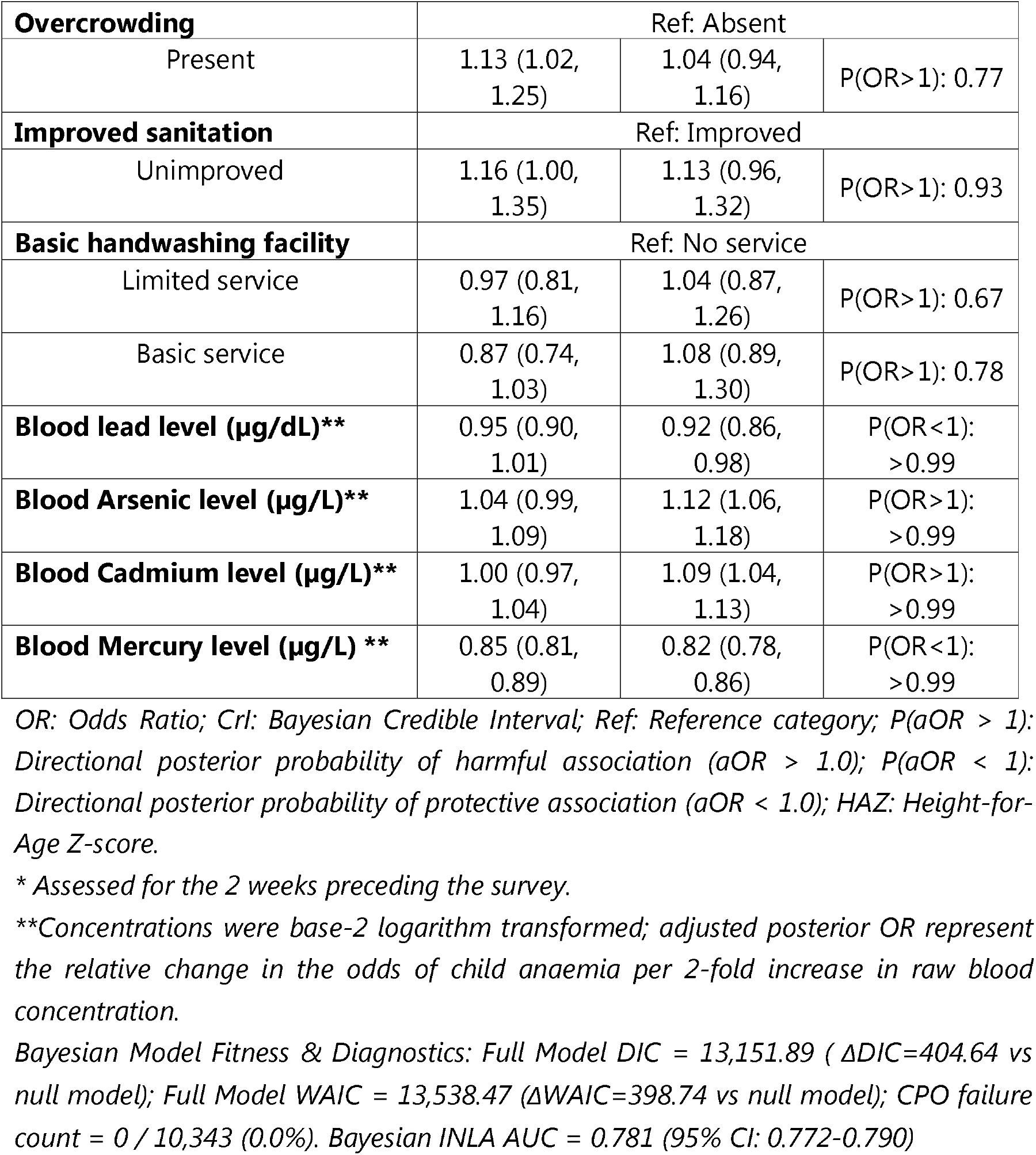
Bayesian multiple logistic regression of factors associated with childhood anaemia among children aged 12-59 months in Bangladesh.

Older child age (aOR = 0.69 per 13.2 months increase, 95% CrI: 0.65-0.72; P(aOR < 1) > 0.99), higher HAZ-score (aOR = 0.88 per WHO Z-score unit, 95% CrI: 0.84-0.91; P(aOR < 1) > 0.99), and female sex (aOR = 0.90, 95% CrI: 0.82-0.98; P(aOR < 1) = 0.99) were associated with lower odds of anaemia. Higher maternal education (aOR = 0.69 vs no education, 95% CrI: 0.56-0.86; P(aOR < 1) > 0.99) and living in the wealthiest household quintile (aOR = 0.80 vs poorest, 95% CrI: 0.66-0.97; P(aOR < 1) = 0.99) were likewise associated with lower anaemia odds. Marked regional variation persisted, with children in Sylhet (aOR = 0.35, 95% CrI: 0.26-0.47) and Chattogram (aOR = 0.57, 95% CrI: 0.44-0.72) exhibiting the lowest odds of anaemia relative to Barishal. The model demonstrated decisive goodness-of-fit over the null model (ΔDIC = 404.64; ΔWAIC = 398.74), 0 CPO failures across N = 10,343 and good AUC-ROC (0.78; 95% CI: 0.77-0.79) (Table 2).

In frequentist survey-weighted multiple logistic regression, effect estimates demonstrated strong consistency with the primary Bayesian hierarchical model (Table 3). Blood arsenic concentration was positively associated with higher odds of childhood anaemia (aOR = 1.10, 95% CI: 1.03-1.17). Similarly, blood cadmium concentration was positively associated with higher odds of childhood anaemia (aOR = 1.07, 95% CI: 1.02-1.12), with both effect estimates expressed per two-fold increase in concentration. Conversely, blood mercury (aOR = 0.86, 95% CI: 0.81-0.90) exhibited inverse associations. Older child age (aOR = 0.72 per 13.2 months increase, 95% CI: 0.68-0.76), higher Height-for-Age Z-score (aOR = 0.89 per WHO Z-score unit, 95% CI: 0.85-0.93), maternal higher education (aOR = 0.67 vs no formal education, 95% CI: 0.53-0.85), and residing in the wealthiest household quintile (aOR = 0.80 vs poorest, 95% CI: 0.64-0.99) were associated with lower odds of child anaemia.

**Table 3.** Frequentist multiple logistic regression of factors associated with childhood anaemia among children aged 12-59 months in Bangladesh.

| Predictor variables | aOR | 95% CI |  | p-value |
| --- | --- | --- | --- | --- |
|  |  | Lower | Upper |  |
| **Child's age** (per 13.2 months) | 0.72 | 0.68 | 0.76 | <0.001 |
| **Child's sex** | Ref: Male |  |  |  |
| Female | 0.91 | 0.83 | 1.01 | 0.071 |
| **Height-for-age Z score** | 0.89 | 0.85 | 0.93 | <0.001 |
| **Diarrhoea\*** | Ref: Absent |  |  |  |
| Present | 0.91 | 0.74 | 1.13 | 0.393 |
| **ARI\*** | Ref: Absent |  |  |  |
| Present | 1.00 | 0.74 | 1.35 | 0.980 |
| **Mother's age** (per 6.3 years) | 1.01 | 0.95 | 1.06 | 0.842 |
| **Mother's level of education** | Ref: No formal education |  |  |  |
| Primary | 0.97 | 0.77 | 1.22 | 0.804 |
| Secondary | 0.81 | 0.65 | 1.00 | 0.047 |
| Higher secondary or more | 0.67 | 0.53 | 0.85 | 0.001 |
| **Wealth index** | Ref: Poorest |  |  |  |
| Poor | 0.82 | 0.71 | 0.95 | 0.007 |
| Middle | 0.85 | 0.72 | 1.00 | 0.044 |
| Rich | 0.87 | 0.73 | 1.03 | 0.110 |
| Richest | 0.80 | 0.64 | 0.99 | 0.038 |
| **Residence** | Ref: Rural |  |  |  |
| Urban | 1.06 | 0.94 | 1.21 | 0.340 |
| **Division** | Ref: Barishal |  |  |  |
| Chattogram | 0.56 | 0.46 | 0.69 | <0.001 |
| Dhaka | 0.60 | 0.49 | 0.74 | <0.001 |
| Khulna | 0.80 | 0.65 | 0.99 | 0.039 |
| Mymensingh | 0.67 | 0.51 | 0.88 | 0.004 |
| Rajshahi | 0.84 | 0.67 | 1.05 | 0.122 |
| Rangpur | 0.64 | 0.52 | 0.79 | <0.001 |
| Sylhet | 0.36 | 0.28 | 0.48 | <0.001 |
| **Overcrowding** | Ref: Absent |  |  |  |
| Present | 1.03 | 0.91 | 1.16 | 0.685 |
| **Improved sanitation** | Ref: Improved |  |  |  |
| Unimproved | 1.07 | 0.91 | 1.27 | 0.410 |
| **Basic handwashing facility** | Ref: No service |  |  |  |
| Limited service | 1.05 | 0.87 | 1.28 | 0.586 |
| Basic service | 1.08 | 0.89 | 1.32 | 0.424 |
| **Blood lead level (µg/dL)\*\*** | 0.94 | 0.87 | 1.00 | 0.060 |
| **Blood Arsenic level (µg/L)\*\*** | 1.10 | 1.03 | 1.17 | 0.004 |
| **Blood cadmium level (µg/L)\*\*** | 1.07 | 1.02 | 1.12 | 0.005 |
| **Blood mercury level (µg/L) \*\*** | 0.86 | 0.81 | 0.90 | <0.001 |

aOR: Adjusted Odds Ratio; CI: Confidence Interval; Ref: Reference category; HAZ: Height-for-Age Z-score.
\* Assessed for the 2 weeks preceding the survey.
\*\*Concentrations were base-2 logarithm transformed; posterior OR represent the relative change in the odds of child anaemia per 2-fold increase in raw blood concentration.
Model diagnostics: Calibration was evaluated using the Archer-Lemeshow goodness-of-fit test for survey data ( $p < 0.001$ ; indicating significant calibration departure). Discrimination was evaluated via survey-weighted AUC-ROC = 0.62 (95% CI: 0.61-0.63).

Regional disparities persisted, with children residing in Sylhet (aOR = 0.36, 95% CI: 0.28-0.48) and Chattogram (aOR = 0.56, 95% CI: 0.46-0.69) exhibiting the lowest odds relative to Barishal (Table 3).

In sensitivity analyses evaluating potential departures from log-linearity using restricted cubic splines (Supplementary Table ST1), multivariable adjusted Wald tests confirmed that log-linear specifications were statistically appropriate for blood arsenic (p_non-linear_=0.980), blood cadmium (p_non-linear_=0.121), and blood mercury (p_non-linear_=0.419). In contrast, a statistically significant non-linear relationship was observed for blood lead (p_non-linear_=0.0014). When blood lead was evaluated across international clinical benchmark categories (<3.5 μg/dL as reference), the apparent inverse association observed in linear models was restricted to sub-clinical exposure levels (3.5 to <5.0 μg/dL : aOR=0.86, 95% CI: 0.76-0.97,p=0.014), but progressively attenuated toward the null at higher concentrations and dissipated completely at or above 10.0 μg/dL (aOR=0.98, 95% CI: 0.77-1.24) (Supplementary Table ST1).

Evaluation of socioeconomic inequality demonstrated pronounced pro-rich inequality in child anaemia prevalence across household wealth ranks (Supplementary Table ST2). Both absolute (SII = −12.39 percentage points, 95% CI: −17.62 to −7.17) and relative (RII = 0.76, 95% CI: 0.67 to 0.85) summary measures confirmed lower anaemia prevalence at higher wealth ranks.

Table 4 presents the estimated PAFs for selected factors, representing the estimated proportional change in child anaemia prevalence if the population were shifted to the specified counterfactual target category. The largest PAFs were observed when maternal education was shifted to higher education, 7.40% (95% CrI: 2.52% to 11.84%), and when household wealth was shifted to the richest quintile, 7.07% (95% CrI: 1.43% to 12.85%). Shifting children aged to the ≥24 months category, representing the hypothetical reduction in anaemia if all children were aged ≥24 months, yielded a PAF of 3.73% (95% CrI: 2.72% to 4.76%). Similarly, shifting stunted children to the non-stunted category, representing the hypothetical reduction in anaemia if no children were stunted, yielded a PAF of 3.71% (95% CrI: 2.50% to 4.86%). Shifting blood cadmium concentrations to ≤0.35 µg/L, representing the hypothetical reduction if all children had concentrations at or below this level, yielded a PAF of 2.37% (95% CrI: 0.09% to 4.57%).

**Table 4:** Bayesian population attributable fractions for child anaemia underspecified counterfactual scenarios.

| **Risk Factor / Exposure Domain** | **Counterfactual Optimal Target** | **PAF % (95% CrI)** |
| --- | --- | --- |
| Maternal Education Level | Higher Education | 7.40% (2.52% to 11.84%) |
| Wealth Quintile | Richest Quintile | 7.07% (1.43% to 12.85%) |
| Child Age (< 24 months) | ≥24 months | 3.73% (2.72% to 4.76%) |
| Stunting Status | Not Stunted (HAZ ≥ -2) | 3.71% (2.50% to 4.86%) |
| Blood Cadmium | $\leq 0.35 \mu\text{g/L}$ (≤ Median) | 2.37% (0.09% to 4.57%) |
| Blood Arsenic | $\leq 1.98 \mu\text{g/L}$ (≤ Median) | 1.85% (-0.40% to 4.09%) |
| Blood Lead | $<5.0 \mu\text{g/dL}$ | 0.28% (-1.59% to 2.10%) |
| Blood Mercury | $\leq 1.52 \mu\text{g/L}$ (≤ Median) | -5.68% (-7.80% to -3.63%) |
| Maternal Age Category | 20–35 years | 1.83% (0.79% to 2.90%) |
| Place of Residence | Urban | -0.07% (-4.23% to 3.90%) |

*PAF, population attributable fraction;*
*CrI, credible interval;*
*HAZ, height-for-age Z-score.*
*PAFs were estimated using counterfactual predictions from the multiple Bayesian* *model and represent the proportional change in predicted anaemia prevalence if each* *exposure were shifted to the specified target while other covariates remained* *unchanged.*
*Positive and negative values indicate lower and higher predicted prevalence,* *respectively.*
*These model-based estimates do not imply causal effects of interventions. Child age* *and residence were included as counterfactual benchmarks rather than modifiable* *targets.*

## Discussion

In this nationally representative study of young children in Bangladesh, nearly half of all children aged 12-59 months were affected by anaemia. This high national prevalence demonstrates that childhood anaemia remains a pervasive public health challenge despite decades of targeted nutritional supplementation and deworming initiatives ^1,13^. Crucially, beyond established demographic and socioeconomic correlates, our study provides novel population-level biomonitoring evidence that environmental heavy metal exposures—specifically blood cadmium and blood arsenic—are independently associated with elevated odds of childhood anaemia in Bangladesh. Furthermore, marked pro-rich socioeconomic inequalities persist, with children from the least wealthy households bearing a disproportionate absolute and relative burden of anaemia compared with their wealthiest peers.

The observed positive association between blood cadmium concentration and childhood anaemia highlights an underrecognized environmental correlate of impaired erythropoiesis in low- and middle-income countries (LMICs). Cadmium is a toxic heavy metal that accumulates in agricultural soils through phosphate fertilizer application, industrial wastewater irrigation, and electronic waste disposal, subsequently bioaccumulating in staple crops such as rice and leafy vegetables ^9^. The ingested cadmium accumulates through absorption at the intestinal brush border, where cadmium competes with inorganic iron for transport via divalent metal transporter-1 (DMT1), impairing systemic iron uptake ^9^. Once absorbed, chronic cadmium accumulation in renal proximal tubular cells can disrupt erythropoietin synthesis through localized nephrotoxicity, while cadmium-induced systemic oxidative stress increases erythrocyte membrane fragility, accelerating premature haemolysis ^9^. The observed association between blood arsenic and child anaemia is consistent with the recognised public health importance of arsenic exposure in Bangladesh, where groundwater contamination represents a major potential exposure source ^12^. Chronic inorganic arsenic exposure impairs erythropoiesis by suppressing bone marrow erythroid progenitor differentiation and promoting low-grade systemic inflammation ^6,7^.

Conversely, higher blood mercury concentration demonstrated an inverse association with childhood anaemia. This phenomenon aligns with the well-documented nutritional confounding or fish paradox frequently described in environmental epidemiology ^35–37^. In non-occupational populations, blood methylmercury functions primarily as a surrogate biomarker for fish consumption ^37^.

In Bangladesh, fish consumption has also been positively associated with mercury concentrations in human hair, supporting dietary fish intake as an important determinant of mercury exposure in the population ^38^. Freshwater and marine fish are major dietary staples that provide bioavailable iron, high-quality animal protein, zinc, selenium, and essential fatty acids that actively support erythropoiesis ^10^. There may be several factors that can contribute to the inverse association between blood mercury and childhood anaemia. One possibility is that higher blood mercury concentrations partly reflect greater fish consumption ^38^, with the nutritional benefits of fish potentially outweighing or confounding any adverse association of mercury exposure with haemoglobin status. Another scenario can be that an increase in RBC mass or hemoglobin density reflects more mercury is present per unit volume, demonstrating a positive correlation ^35,39,40^. In contrast to bioaccumulative agricultural and industrial contaminants such as cadmium and groundwater arsenic, mercury exposure in this setting primarily tracks animal-source food intake. Blood lead concentration showed a modest inverse association in the primary linear models, but sensitivity analyses indicated significant non-linearity. The inverse association was observed only at 3.5 to <5.0 μg/dL, while associations at 5.0 to <10.0 μg/dL and ≥10.0 μg/dL were not statistically significant. This pattern may similarly reflect complex co-exposure dynamics, non-linear dose-response thresholds, or unmeasured nutritional buffer mechanisms ^5,8^.

Socioeconomic disparities remain central to the structural distribution of childhood anaemia. Anaemia was more prevalent among children from poorer households and those whose mothers had lower educational attainment. These socioeconomic differences may reflect multiple unmeasured pathways associated with household disadvantage, although the present analysis could not directly evaluate dietary quality, environmental exposures, or healthcare access ^1,4^. The counterfactual PAF analysis provided an additional population-level perspective on the regression findings. It highlighted which factors were associated with the largest differences in predicted anaemia prevalence. The results indicate that maternal education and household wealth were associated with the largest differences in predicted population anaemia prevalence. Younger age and lower linear growth Z-scores were also associated with higher odds of anaemia, highlighting the vulnerability of younger and stunted children. Blood cadmium was associated with a smaller, but positive, model-based difference in predicted anaemia prevalence, whereas the corresponding estimates for arsenic and lead were close to or included the null.

This study has several strengths, including the use of the first nationally representative South Asian survey incorporating simultaneous four-metal blood biomonitoring alongside haemoglobin measurement in over 10,000 children. The integration of complex survey weights with Bayesian hierarchical modelling allowed the analysis to account for community-level clustering through PSU random intercepts. Model diagnostics showed no CPO failures, indicating no CPO-based numerical or predictive instability.

Several limitations should be noted. The cross-sectional study design precludes temporal sequence and causal inference. Additionally, heavy metal biomonitoring was restricted to a sub-sample of survey participants, although potential selection bias was minimized through normalized sampling weights. The survey documentation reported analytical precision concerns for arsenic, cadmium, and mercury. Therefore, associations involving these biomarkers, including the corresponding model-based counterfactual PAF estimates, should be interpreted cautiously, as measurement imprecision may have affected the estimated associations. Restricted cubic splines identified non-linear blood lead dose–response patterns, but the cross-sectional design limited assessment of temporal toxicant accumulation. Finally, the survey lacked iron biomarkers, such as serum ferritin and soluble transferrin receptor, and genetic screening for inherited haemoglobinopathies. These limitations prevented differentiation of iron-deficiency, inflammatory, and genetic causes of anaemia.

## Conclusion

Childhood anaemia in Bangladesh is associated with socioeconomic disadvantage, lower linear growth, structural inequality, and environmental toxicant exposure.

Progress towards SDG 2.2 and the WHO Global Nutrition Targets may benefit from broadening national anaemia strategies beyond iron supplementation alone. The findings highlight the potential importance of integrating nutritional, environmental, and socioeconomic considerations, including environmental monitoring and efforts to address underlying social inequalities. Multisectoral collaboration across public health, agriculture, and environmental sectors may help inform more comprehensive strategies to address the population burden of childhood anaemia.

## Data Availability

All data produced are available online at: https://mics.unicef.org/

## Acknowledgements

All the authors work at icddr,b. icddr,b is grateful to the Government of Bangladesh and Canada providing core/unrestricted support.

## Author contributions

MFAF, RR, TA and MM conceived and designed the study. MRI, MFAF and AKR were associated with data collection, curation, analysis and interpretation. MFAF, MRI and AKR wrote the manuscript. RR, TA and MM critically reviewed and edited the manuscript. All the author(s) read and approved the final manuscript for submission.

## Ethical Standards Disclosure

Current manuscript used data from the publicly available Multiple Indicator Cluster Survey (MICS) Bangladesh 2025 data. As this was a secondary data analysis, not ethical clearance was required. The Bangladesh MICS 2025 survey protocol, including the blood specimen collection procedure, was formally reviewed and approved by the icddr,b Ethical Review Committee in November 2024 (Protocol Reference: PR-24113). Written informed consent was obtained from mothers or legal guardians prior to blood collection. Participants identified with severe anaemia or dangerously elevated heavy metal concentrations were provided immediate counselling and referred through the Ministry of Health and Family Welfare to appropriate medical facilities for clinical management. Anonymized microdata was accessed for research purposes in accordance with BBS and UNICEF data sharing policies.

## Availability of data

Data pertaining to this manuscript can be obtained upon request from https://mics.unicef.org/

## Declaration of AI usage

This work has benefited from the use of artificial intelligence (AI) tools to support the writing and editing process. Specifically, OpenAI’s ChatGPT (GPT-5.6) was used to assist in language refinement, organisation of content, and enhancement of clarity and academic tone. All substantive ideas, data interpretation, and critical analysis are the original work of the authors. The AI was used solely as a linguistic and editorial aid, and its contributions were reviewed and approved by the authors to ensure accuracy and scholarly integrity.

