## Supplementar Table ST1, Supplementary Table ST2 for "Environmental heavy metal exposure, socioeconomic inequalities, and childhood anaemia in Bangladesh: a nationally representative, survey-weighted Bayesian hierarchical analysis"

**Supplementary Table ST1. Sensitivity analyses of non-linear dose–response relationships and clinical threshold categories for blood heavy metals in relation to childhood anaemia**

| **Heavy Metal Biomarker** | **Frequentist Model aOR (95% CI)*** | **Non-Linearity p-value** | **Functional Form** |
| --- | --- | --- | --- |
| Blood Arsenic (μg/L) | 1.10 (1.03, 1.17) | 0.980 | Log-linear |
| Blood Cadmium (μg/L) | 1.07 (1.02, 1.12) | 0.121 | Log-linear |
| Blood Mercury (μg/L) | 0.86 (0.81, 0.90) | 0.419 | Log-linear |
| Blood Lead (μg/dL) | 0.94 (0.87, 1.00) | 0.0014 | Non-linear |
| **Blood Lead Category** | | | |
| **Categories** | **Frequency (%)*** | **aOR (95% CI)** | **p-value** |
| <3.5 μg/dL | 4112 (39.8%) | Ref. | Ref. |
| 3.5 to <5.0 μg/dL | 2947 (28.5%) | 0.86 (0.70-0.97) | 0.014 |
| 5.0 to <10.0 μg/dL | 2782 (26.9%) | 0.89 (0.78-1.01) | 0.076 |
| ≥10.0 μg/dL | 502 (4.8%) | 0.98 (0.77-1.24) | 0.849 |

*aOR: adjusted Odds Ratio; CI: Confidence Interval; df: degrees of freedom.*

*All models were estimated using survey-weighted logistic regression. All models were mutually adjusted for child age, sex, height-for-age Z-score, diarrhoea, acute respiratory infection, maternal age, maternal education, household wealth quintile, urban/rural residence, administrative division, household overcrowding, improved sanitation, basic handwashing facility, and concurrent blood concentrations of the other three heavy metals.*

**Model evaluates continuous log2-transformed blood metal concentrations (odds ratio per 2-fold increase).*

***Unweighted n and survey-weighted percentages were reported*

**Supplementary Table ST2. Health Inequality Summary Measures for Child Anaemia Across Household Wealth Quintiles**

| **Inequality Indicator** | **Estimate (95% CI)** |
| --- | --- |
| SII | -12.39 (-17.62 to -7.17) |
| RII | 0.76 (0.67 to 0.85) |

*SII: Slope Index of Inequality; RII: Relative Index of Inequality; CI: Confidence Interval*
